# Class imbalance correction in artificial intelligence models leads to miscalibrated clinical predictions: a real-world evaluation

**DOI:** 10.64898/2026.03.04.26347634

**Authors:** Mathias Roesler, Cameron Wells, Gabriel Schamberg, Junyi Gao, Luke Boyle, Ewen Harrison, Greg O’Grady, Chris Varghese

## Abstract

**Background:** Predictive models employing machine learning algorithms are increasingly being used in clinical decision making, and improperly calibrated models can result in systematic harm. We sought to investigate the impact of class imbalance correction, a commonly applied preprocessing step in machine learning model development, on calibration and modelled clinical decision making in a large real-world context.

**Methods:** A histogram boosted gradient classifier was trained on a highly imbalanced national dataset of *>*1.8 million patients undergoing surgery, to predict the risk of 90-day mortality and complications after surgery. Class imbalance correction strategies including random oversampling, synthetic minority oversampling technique, random under-sampling, and cost-sensitive learning were compared to the natural distribution (‘natural’). Models were tested and compared with classification metrics, calibration plots, decision curve analysis, and simulated clinical impact analysis.

**Results:** The natural model demonstrated high performance (AUROC 0.94, 95% CI 0.94–0.95 for mortality; 0.84, 95% CI 0.84–0.85 for complications) and calibration (log loss 0.05, 95% CI 0.04–0.05 for mortality; 0.23, 95% CI 0.23–0.24 for complications). Class imbalance mitigation (CSL, ROS, RUS, and SMOTE) did not improve AUROC or AUPRC but increased recall and F_1_ scores at the expense of precision and accuracy. However, these methods severely compromised model calibration, leading to significant over-prediction of risks (up to a 62.8 % increase) as further evidenced by increased log loss across all mitigation techniques. Decision curve analysis and clinical scenario testing confirmed that the natural model provided the highest net benefit.

**Conclusion:** Class imbalance correction methods result in significant miscalibration, leading to possible harm when used for clinical decision making.

## INTRODUCTION

Artificial intelligence (AI) is increasingly being applied in the development of clinical prediction models in healthcare ^1;2;3^. Common architectures deployed on tabular medical data for risk prediction include random forests, gradient boosting methods (*e*.*g*., XGBoost), and deep neural networks. The improved performance and flexibility of these approaches has seen widely used clinical prediction models transition from traditional regression-based models to ML alternatives; with these alternatives able to provide precise individualised probabilities for various clinical outcomes ^4^.

Notably, however, a frequent challenge in developing clinical risk prediction models in medical settings is class imbalance ^5;6^. Accordingly, when training machine learning (ML) models to predict such rare events, there may be a concern that the models will learn a bias in favour of the dominant class. To address this concern, several strategies are commonly applied to address class imbalance ^7^, including data-level strategies such as under sampling the majority class ^8^, oversampling the minority class ^8^, and synthetic minority oversampling techniques (SMOTE) ^9^; or algorithm-level strategies such as cost-sensitive learning, and model-level penalties for minority event misclassification ^10^. These methods are summarised in Table 1, with comprehensive reviews of class imbalance mitigation methods previously been published ^11;12^.

**Table 1.**
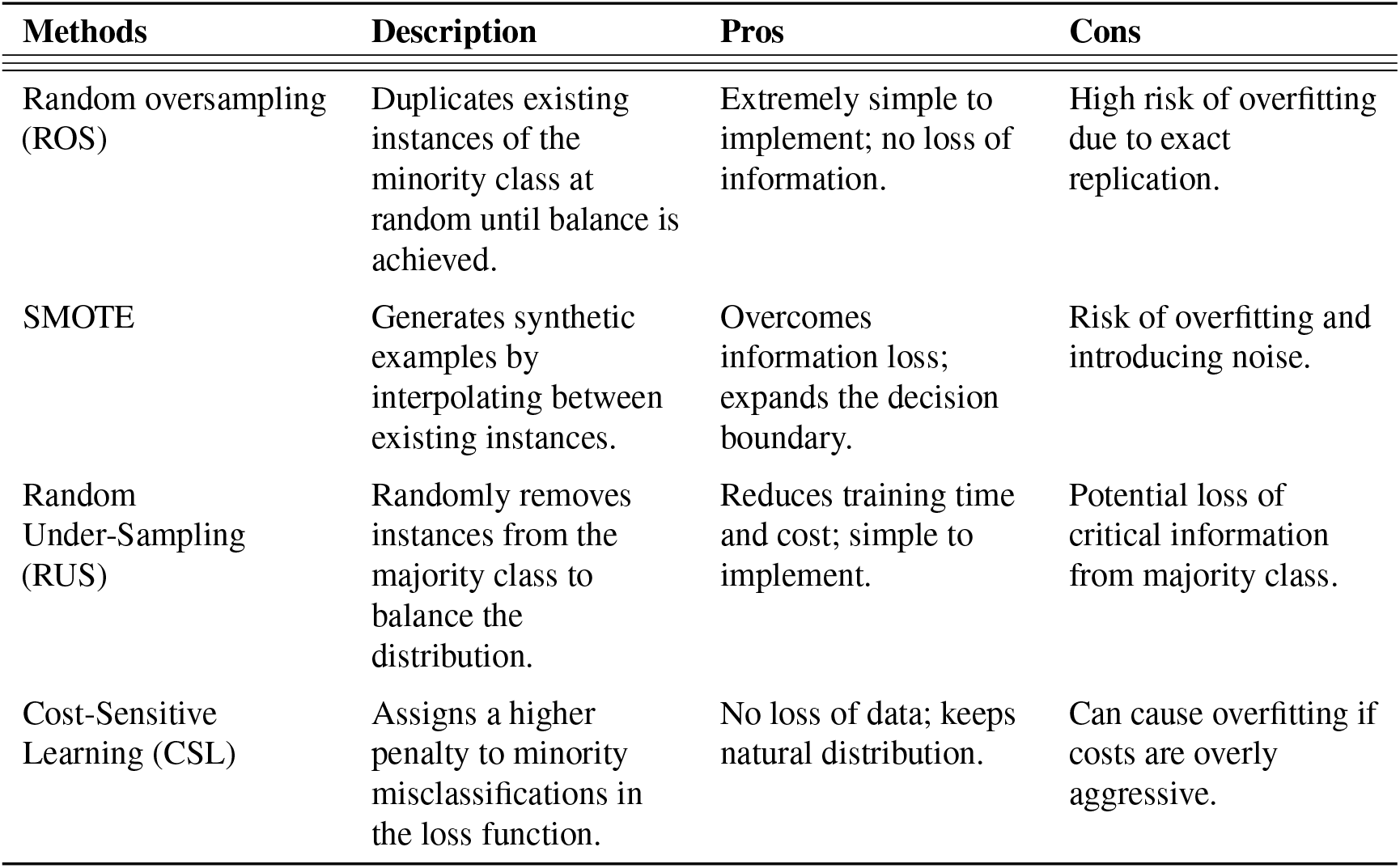
Overview of Class Imbalance Mitigation Methods.

| Methods | Description | Pros | Cons |
| --- | --- | --- | --- |
| Random oversampling (ROS) | Duplicates existing instances of the minority class at random until balance is achieved. | Extremely simple to implement; no loss of information. | High risk of overfitting due to exact replication. |
| SMOTE | Generates synthetic examples by interpolating between existing instances. | Overcomes information loss; expands the decision boundary. | Risk of overfitting and introducing noise. |
| Random Under-Sampling (RUS) | Randomly removes instances from the majority class to balance the distribution. | Reduces training time and cost; simple to implement. | Potential loss of critical information from majority class. |
| Cost-Sensitive Learning (CSL) | Assigns a higher penalty to minority misclassifications in the loss function. | No loss of data; keeps natural distribution. | Can cause overfitting if costs are overly aggressive. |

Class imbalance is particularly common in surgical settings, with perioperative risk stratification tools typically targeting infrequent critical events such as postoperative mortality or complications. Widely used surgical risk stratification tools include the American College of Surgeons National Surgical Quality Improvement Program (NSQIP) Risk Calculator in the United States ^4^, the National Emergency Laparotomy Audit (NELA) model developed in the United Kingdom for emergency laparotomy ^13^, and NZRisk in New Zealand ^14^. Predictions from such surgical risk stratification tools are useful in guiding patient discussions, obtaining informed consent, and allowing needs-based resource allocation. Such tools are particularly useful in high-risk and emergency settings to guide clinical decision making ^15;16^. Surgical risk prediction models therefore present a valuable scenario where outcomes of interest for prediction are infrequent, and hence class imbalance mitigation strategies may be commonly applied.

Here, we present a real-world comparison of class imbalance mitigation strategies when applied to a large national cohort of patients undergoing surgery. This cohort is characterized by a low incidence of outcomes of the outcomes being predicted, namely, death (*<*2%) or postoperative complications (*<*15%) occurring within 90 days after surgery. Although complication rates vary according to the combination of cases in the surgical cohort examined ^14;17^, this cohort is representative of the public healthcare system that provides surgical care to most of the nation and is representative of the typical national samples used to train previously published surgical risk stratification tools ^14;4^. In this real-world national surgical cohort, we aimed to assess the impact of class imbalance mitigation strategies on model performance, calibration, and clinical decision making.

## MATERIALS AND METHODS

This study was conducted following the TRIPOD+AI guidelines ^18^.

### Data

This was a national, population-based retrospective sample of adults undergoing surgery under general or neuraxial anaesthesia in public hospitals in Aotearoa New Zealand between 1 January 2010 and 31 December 2024. Ethical approval was obtained from the Auckland Health Research Ethics Committee (AH29056).

For each patient, 12 distinct features were extracted to serve as model inputs (see Table 4 in Supplementary Materials).

The primary outcomes of interest were 90-day mortality and the occurrence of any postoperative complications at 90 days (further detailed in Table 2).

**Table 2.** Clinical outcomes.

| Outcome | Train set<br>(2010 – 2022)<br>N=1,867,300 (87 %) | Test set<br>(2023 – 2024)<br>N=287,123 (13 %) |
| --- | --- | --- |
| **90-day mortality, count (%)** |  |  |
| Yes | 24,990 (1.3) | 4,406 (1.5) |
| No | 1,842,310 (98.7) | 282,717 (98.5) |
| **Any complications, count (%)** |  |  |
| Yes | 175,386 (9.4) | 31,558 (11.0) |
| No | 1,691,914 (90.6) | 255,565 (89.0) |

A total of 6,378 patients were removed from the dataset because information from at least one of the features was missing.

### Data processing

Several preprocessing operations were performed on the data. First, all categorical variables were transformed using the OrdinalEncoder from the Python package scikit-learn, mapping qualitative features into a numerical format. Following this, numerical features were scaled by applying the StandardScaler from the Python package scikit-learn to all non-binary features. Finally, the distribution of the M3 score ^19^ was addressed by placing values into five bins and capping them at a maximum of 4. This operation helped mitigate the impact of outliers that could potentially skew the learning process. The full list of features and the data distributions among the different sets are presented in the supplementary Table 4.

### Model training

A histogram gradient boosting (HGB) model was used, matching the current NSQIP model ^4^. The HGB model was implemented using the scikit-learn Python library. The model discretized continuous features into a maximum of 255 bins to accelerate the splitting process. The architecture was optimised using a leaf-wise growth strategy with a maximum of 50 leaves per tree and a minimum of 100 samples per leaf. The model was trained for a maximum of 200 iterations with early stopping if the loss did not improve for 10 consecutive iterations. The learning rate was 0.05 and no L2 regularisation was used.

### Model validation

A temporal cross-validation was implemented using year of surgery. Data from 2010 to 2022 formed the primary training and validation development set, representing 87% of the full dataset. The years 2023 and 2024 were reserved as an independent unseen test set to evaluate final model performance, and represented 13% of the total dataset (see Figure 6).

### Class imbalance

The natural model was trained on the entire dataset with the natural distribution. Other models were trained with class imbalance mitigation methods applied ^8;10;9^. In this paper, four common methods were tested, namely one algorithmic approach: cost-sensitive learning (CSL) and three sampling approaches: random oversampling (ROS), random under-sampling (RUS), and Synthetic Minority oversampling Technique (SMOTE). A summary of the different methods along with some of the advantages and drawbacks of each method are presented in Table 1. All sampling procedures were applied strictly within the training folds to ensure class distributions were perfectly balanced across experimental groups while preventing data leak to the test set.

In additional analyses, sampling ratios were varied for the three sampling-based methods (ROS, RUS, and SMOTE), to evaluate the impact of various degrees of class imbalance correction. The imbalance ratio (imbalance ratio) ^10^ for a dataset with a binary prediction label was defined as:

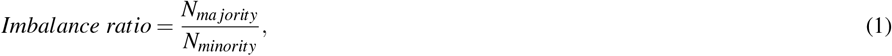

where *N*_*majority*_ and *N*_*minority*_ are the number of samples in the majority and minority classes, respectively. Therefore, *imbalanceratio* ∈[1, ∞), where 1 represents a perfectly balanced dataset and imbalance ratio increases as the number of minority samples in the dataset decreases. Thus, there are two possibilities to reduce the imbalance ratio:

- decrease *N*_*majority*_ by removing examples belonging to the majority class - *e*.*g*., through methods such as RUS.
- increase *N*_*minority*_ by adding examples to the minority class - *e*.*g*., ROS or synthetic additions with SMOTE.

In this paper, the imbalance ratio for the 90-day mortality and the complications labels were 73.2 and 9.6, respectively. Four different imbalance ratios were tested for each label and each sampling method at 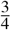, 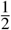, and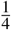of the natural imbalance ratio and a perfectly balanced set with an imbalance ratio of 1.

### Evaluation metrics

The evaluation metrics selected to evaluate the models were selected based on predictive model development guidelines ^20;1^.

Discrimination was assessed using area under the receiver operating characteristic (AUROC) curve, and the area under the precision-recall (AUPRC) curve. Calibration was assessed by the logarithmic loss (log loss), defined as:

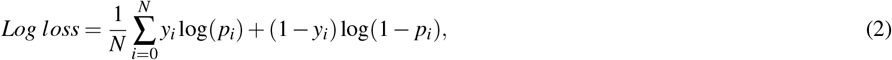

where N is the total number of samples, *y*_*i*_ is the i^th^ binary label (1 or 0), and *p*_*i*_ is the predicted probability of the i^th^ label between 1 and 0.

The log loss was chosen to evaluate model calibration, as its logarithmic penalty is more sensitive to errors near the probability extremes (0 and 1) compared to the quadratic penalty of the Brier Score, making it a robust indicator of predictive certainty in skewed distributions ^21^.

We also report accuracy, precision, recall, and F_1_ score were also calculated with a decision threshold set to 0.5, as is commonly reported in ML papers. It is worth noting that these metrics depend on the classification threshold chosen and as such can be artificially inflated if threshold at a probability significantly different to the pretest probability. All metrics values are presented as the mean from all the training folds with a 95% confidence interval.

Predicted probability distributions and calibration curves (reliability diagrams) were also generated for each model. For the sampling methods (ROS, RUS, and SMOTE) the metrics, calibration curves, and the predicted probability distributions were also calculated at different imbalance ratios. When applicable, the 95% confidence intervals were calculated via bootstrapping with 1000 iterations. Finally, to assess the impact of class imbalance mitigation in a clinical setting, decision curve analysis (DCA) graphs were made for the model without class imbalance mitigation, the model with CSL, and the models using balanced sets from ROS, RUS, and SMOTE methods. DCA measures the clinical value of models by assessing the net benefit over a continuous range of thresholds ^22^. This approach accounts for the inherent trade-off between the benefits of identifying true positives and the harms of false positives. The net benefit is defined as:

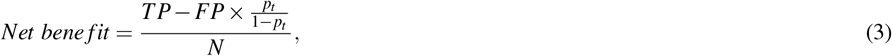

where N is the total number of samples, TP the true positive count, FP the false positive count, and *p*_*t*_ the probability threshold. The ratio 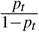 serves as a weighting factor based on the relative harm of a false positive intervention.

Additionally, a hypothetical scenario was used to assess the impact of class imbalance mitigation-related effects on model output probabilities on clinical decision making. Here, we modelled the error in predicted probabilities and the possible perpetuation of clinical errors if made at a set decision threshold. For this scenario, an arbitrary risk threshold of *<*10 % for postoperative complications and *<*2 % for postoperative mortality was used where clinical decisions based on these probabilities were being made.

## RESULTS

The training cohort included 1,867,300 patients (53.5% female; median age 51±31 years) with a 90-day mortality rate of 1.3% and 90-day any complication rate of 9.4%. The test cohort included 287,123 patients (54.2% female; median age 54±32 years) with a 90-day mortality rate of 1.5% and 90-day any complication rate of 11.0%. See Table 2 for the details about the outcomes and Table 4 for the cohort information.

### Model calibration and evaluation

Figure 1 shows the evaluation metrics (Figure 1A and 1C) and calibration curves (Figure 1B and 1D) for the natural model, the model using CSL and the models trained on balanced datasets (using ROS, RUS, and SMOTE). In both cases, the AUROC and the AUPRC were unchanged regardless of the method used to address class imbalance with respective mean values and standard deviations across methods of 0.94±0.00 and 0.19 ±0.01 for the 90-day mortality outcome and 0.84±0.00 and 0.41±0.01 for the complications outcome. The accuracy and precision decreased when using a class imbalance mitigation method, however, the F_1_ and recall showed improvements as expected in being a thresholded at 50% dependent metric.

**Figure 1:**
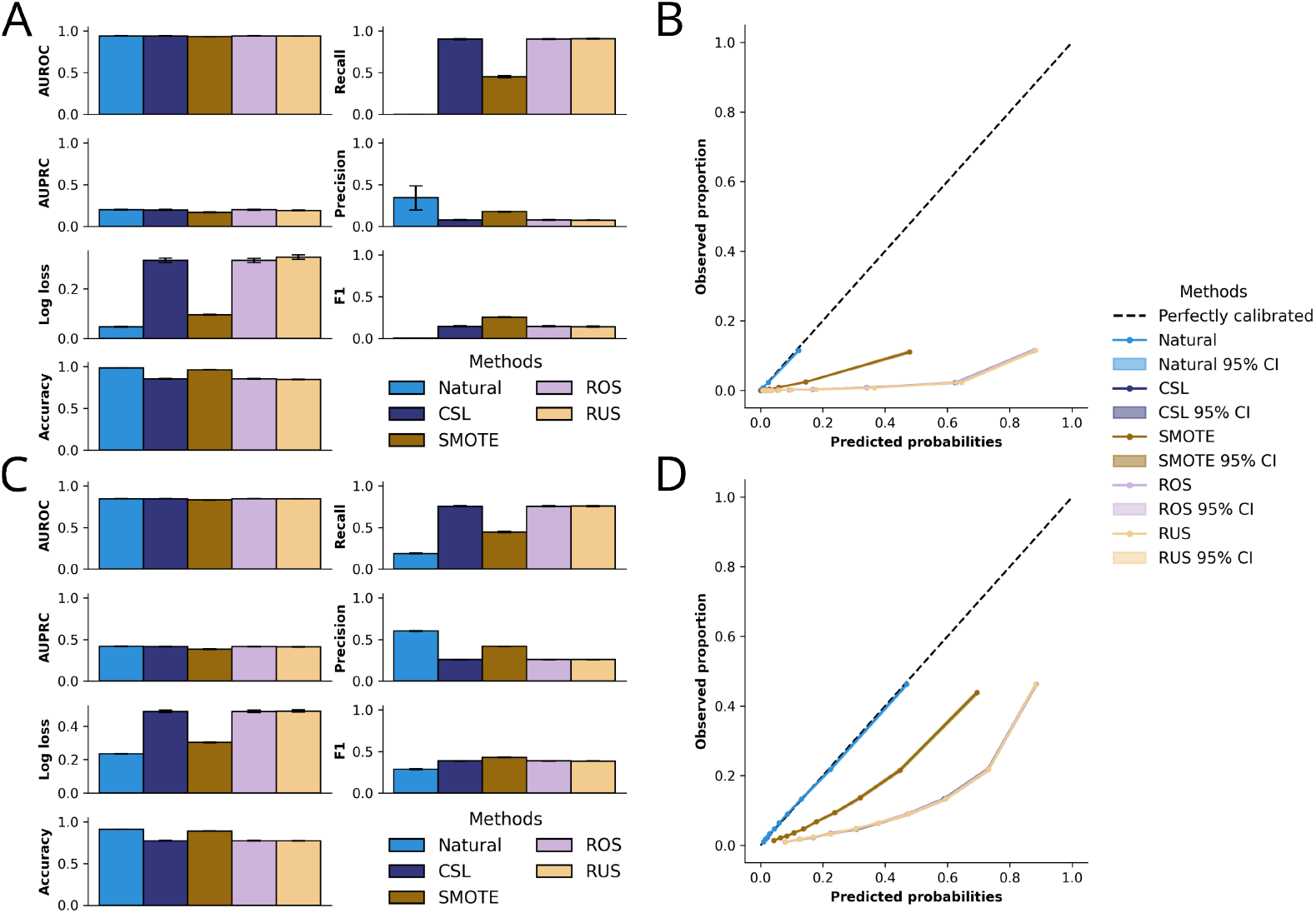
Evaluation metrics and calibration curves for the natural model, the model using CSL and the models trained on balanced datasets using the sampling methods (ROS, RUS, and SMOTE). Panels A and B show the results for the 90-day mortality label and panels C and D the ones for the complications label. Class imbalance mitigation methods improved the recall and F_1_ score but negatively impacted the calibration and log loss. The AUROC and AUPRC, however, were unaffected.

The log loss was lowest for the natural model at 0.05, 95% CI [0.04, 0.05] and 0.23, 95% CI [0.23, 0.24], for the 90-day mortality the complications outcomes, respectively. Notably however, the log loss substantially increased (indicating worse calibration) for all models in which class imbalance mitigation methods were applied. The SMOTE method showed a smaller increase at 0.10, 95% CI [0.09, 0.10] and 0.30, 95% CI [0.29, 0.30] for the 90-day mortality the complications outcome, respectively. The CSL, ROS and RUS methods had similar log loss values at 0.32, 95% CI [0.31, 0.32], 0.32, 95% CI [0.31, 0.32] and 0.33, 95% CI [0.32, 0.34], respectively, for the 90-day mortality label and 0.49, 95% CI [0.48, 0.50], 0.49, 95% CI [0.48, 0.50] and 0.49, 95% CI [0.48, 0.50] for the complications label.

The calibration curves, confirm the trends in log loss, with deterioration in model calibration when class imbalance mitigations were applied. The natural model demonstrated the best calibration. Where as all class imbalance correction strategies systematically induced a tendency to over-estimate the risk of both 90-day mortality and complications.

Figure 2 shows the predicted probability distributions of the different models for both outcomes. The baseline model probability distributions for both outcomes were skewed towards 0. This effect, due to the majority label being no mortality or complication, was more pronounced for the 90-day mortality outcome, which had a much higher imbalance ratio (73.2) compared to the complications (9.6). The class imbalance mitigation methods had the same effect on the predicted probabilities, namely shifting the distribution to be more uniform. This effect was influenced by the imbalance ratio, with a greater shift being observed with smaller imbalance ratios. The distribution for the CSL method closely resembled the ROS and RUS distributions with a balanced dataset (imbalance ratio = 1.0).

**Figure 2:**
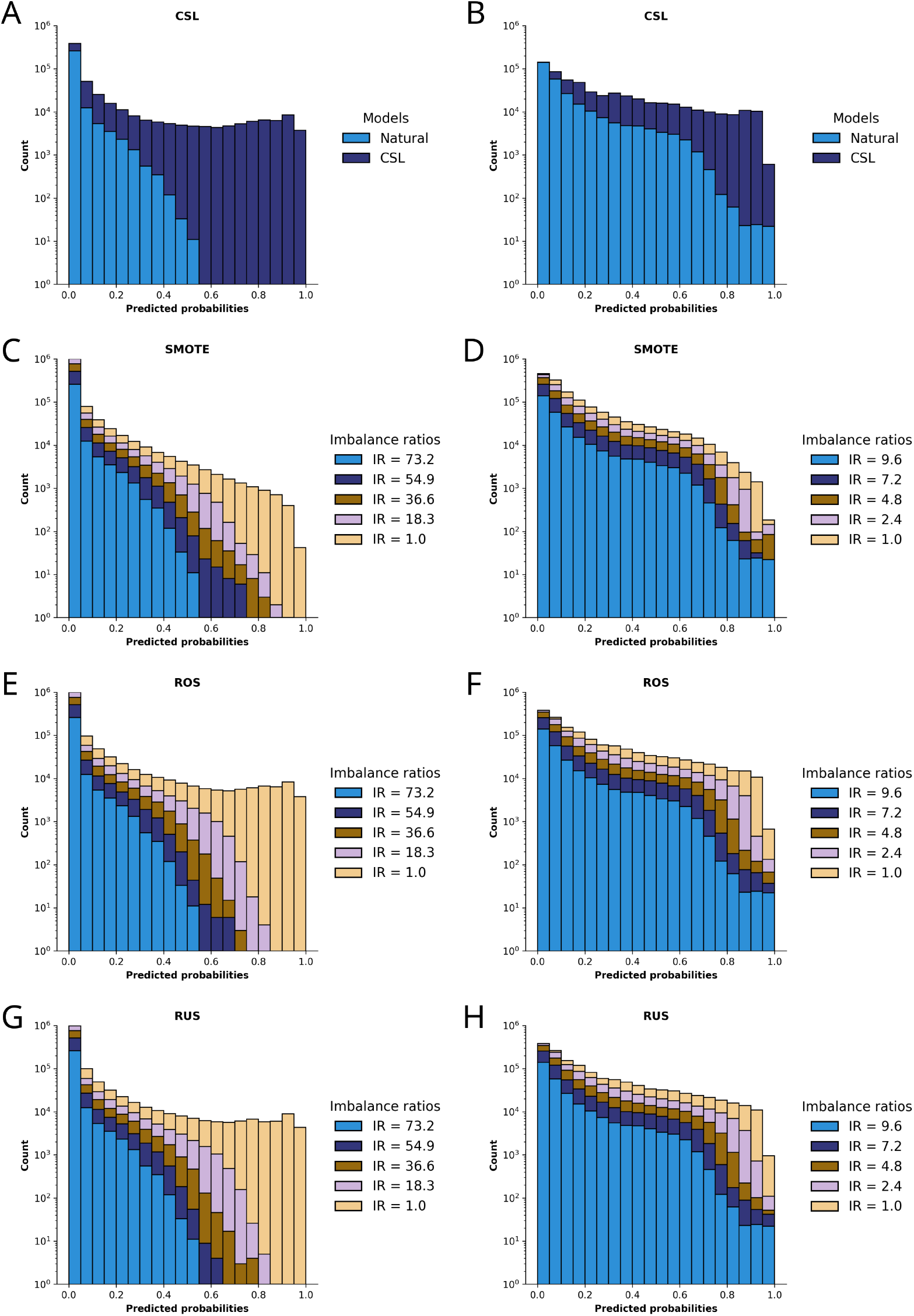
Predicted probability distributions of the different models. The graphs on the left (panels A, C, E, and G) are the distributions for the 90-day mortality label and the ones on the right (panels B, D, F, and H) for the complications label. The probability distributions of the baseline models (imbalance ratio = 73.2 and 9.6 for the 90-day mortality and complications labels, respectively) skewed towards 0, *i*.*e*. the majority label. The class imbalance mitigation methods shifted the distributions to be more uniform.

Figure 3 presents the evaluation metrics for the models using sampling methods (ROS, RUS, and SMOTE) at different imbalance ratios. Similar to Figure 1, the AUROC and AUPRC remained constant for all methods as the imbalance ratio decreased. The accuracy only decreased when the dataset was balanced and the imbalance ratio was 1. Like above, the log loss showed the most significant changes when models were fully balanced. The recall and F_1_ score increased as the imbalance ratio decreased, while the precision decreased, which is expected given these metrics are threshold at 0.5, suiting a balanced training evaluation.

**Figure 3:**
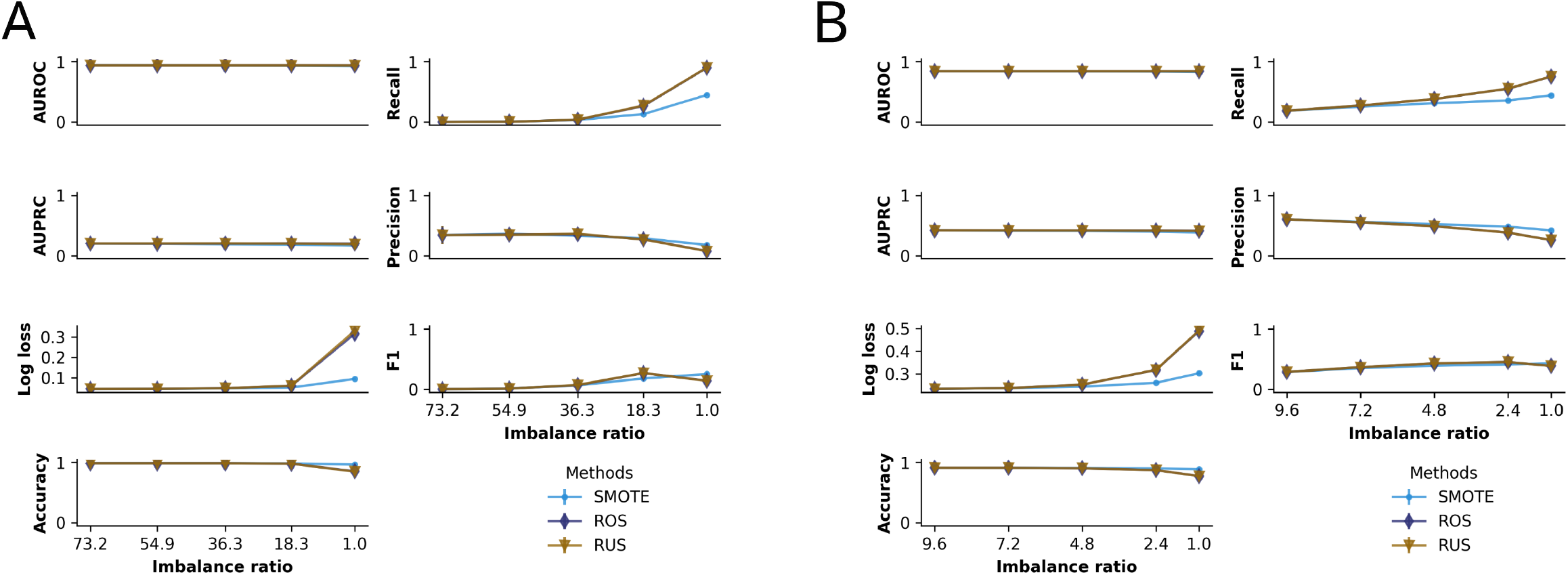
Effect of different imbalance ratios on the evaluation metrics. Panels A and B show the metrics for the three models using sampling methods to address class imbalance, for the 90-day mortality and complications labels, respectively. These results show that perfectly balancing a dataset does not always yield the best models.

Figure 4 shows the calibration curves at different imbalance ratios for the different methods. The three sampling methods (ROS, RUS, and SMOTE) exhibited a proportionally worse calibration as the imbalance ratio was increased. As the imbalance ratio decreased, the curve moved further away from the perfect calibration line toward more systematic over-estimation of risk probabilities for 90-day mortality or any complication.

**Figure 4:**
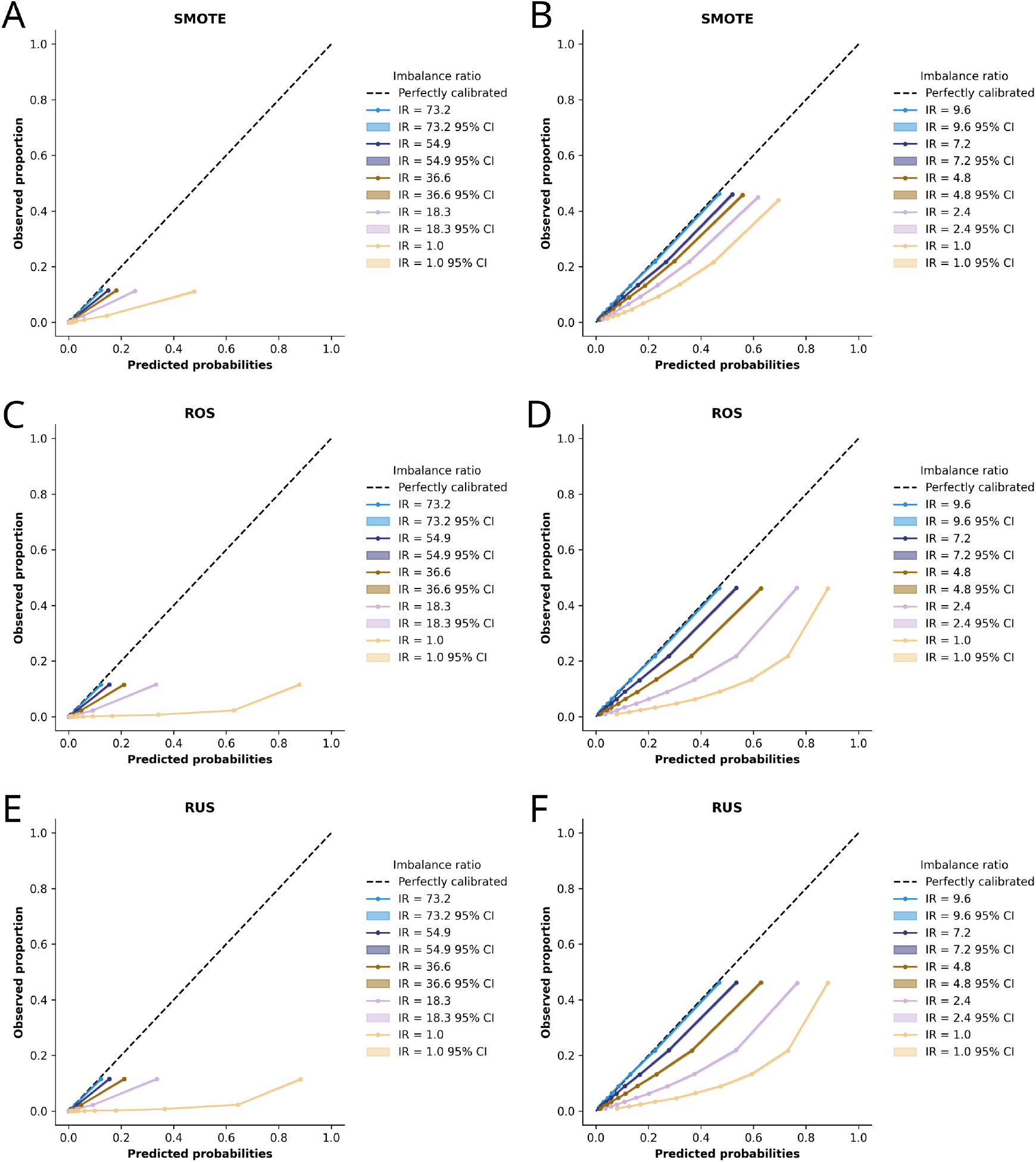
Calibration curves for the different sampling methods with respect to the imbalance ratio. The graphs on the left (panels A, C, and E) show the calibration curves for the 90-day mortality label while those on the right (panels B, D, and F) correspond to the complications label. As the imbalance ratio decreased, the calibrations skewed and the models over-predicted the risk of events occurring.

### Clinical decision making impact analysis

Figure 5 shows the decision curve analysis graphs for the different models and outcomes. DCA clearly demonstrates that for any given threshold (between 0–0.1 for the 90-day mortality label and 0–0.5 for the complications label), the natural model had a higher net benefit compared to any model trained with class imbalance mitigation applied.

**Figure 5:**
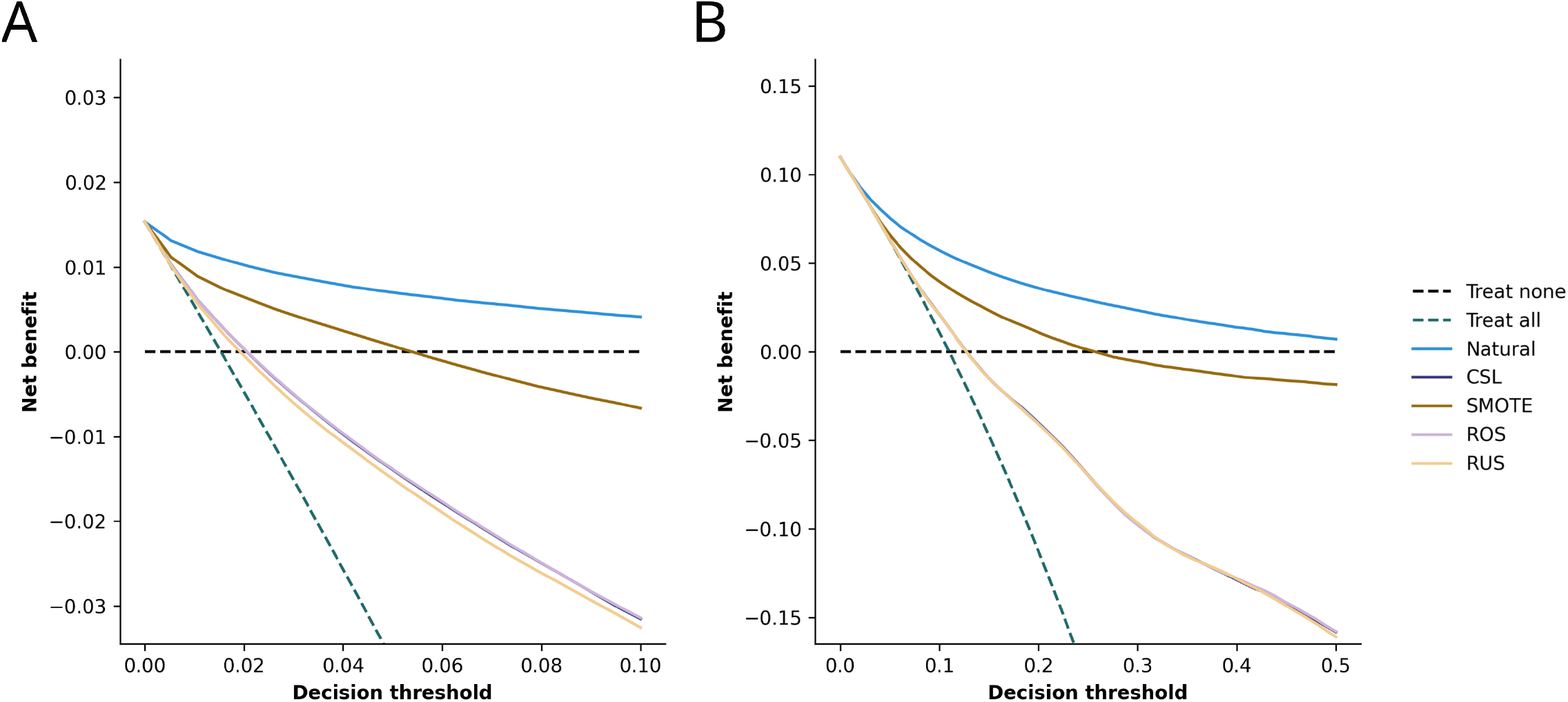
Decision curve analysis graphs for the 90-day mortality label (panel A) and the complications label (panel B). For both labels, the natural model outperformed the models that applied a class imbalance mitigation method with the highest net benefit across all thresholds.

**Figure 6:**
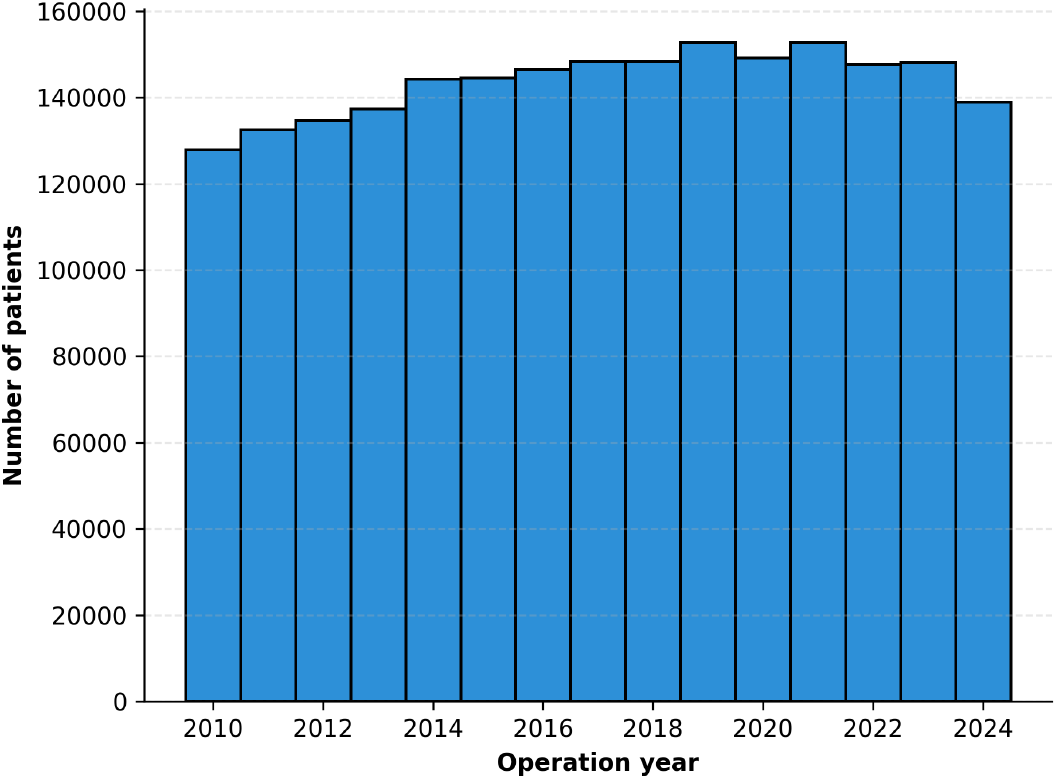
Number of patients per operation year. The distribution showed that the number of patients increased between 2010 and 2014 but remained stable afterwards. This distribution confirms the use of a time-based cross-validation training scheme.

To exemplify the clinical consequences of utilising miscalibrated models, we present a scenario where these models are deployed to a hypothetical clinical settings and contribute to clinical decision making (Table 3). First we take a threshold of *>*2% mortality risk and *>*10% any complication risk, to classify high-risk surgery, at which a different clinical decision may be made (*e*.*g*., preoperative decision to operate or pre-arranged intensive care unit bed). Applying the natural model, with no class imbalance mitigation, 16.1% of surgeries in the test set were predicted to be high-risk of mortality, and 31.0% were at high risk of complications. When class imbalance mitigations were applied, significantly more patients were classified as high-risk, possibly resulting in substantially increased proportion of inappropriate clinical decision making (Table 1).

**Table 3.**
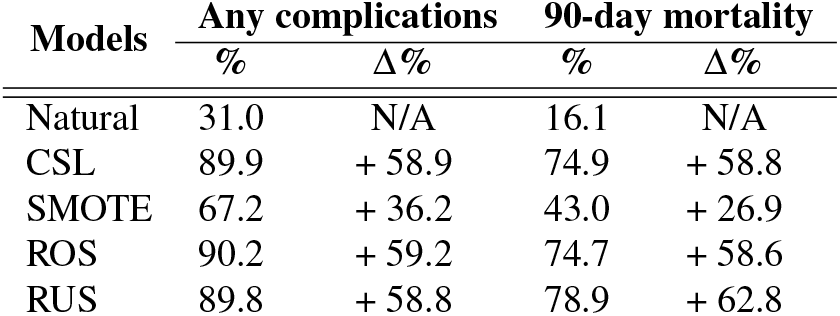
Percentage of risky surgeries from the clinical test scenario.

| Models | Any complications |  | 90-day mortality |  |
| --- | --- | --- | --- | --- |
| | % | $\Delta\%$ | % | $\Delta\%$ |
| Natural | 31.0 | N/A | 16.1 | N/A |
| CSL | 89.9 | + 58.9 | 74.9 | + 58.8 |
| SMOTE | 67.2 | + 36.2 | 43.0 | + 26.9 |
| ROS | 90.2 | + 59.2 | 74.7 | + 58.6 |
| RUS | 89.8 | + 58.8 | 78.9 | + 62.8 |

**Table 4.**
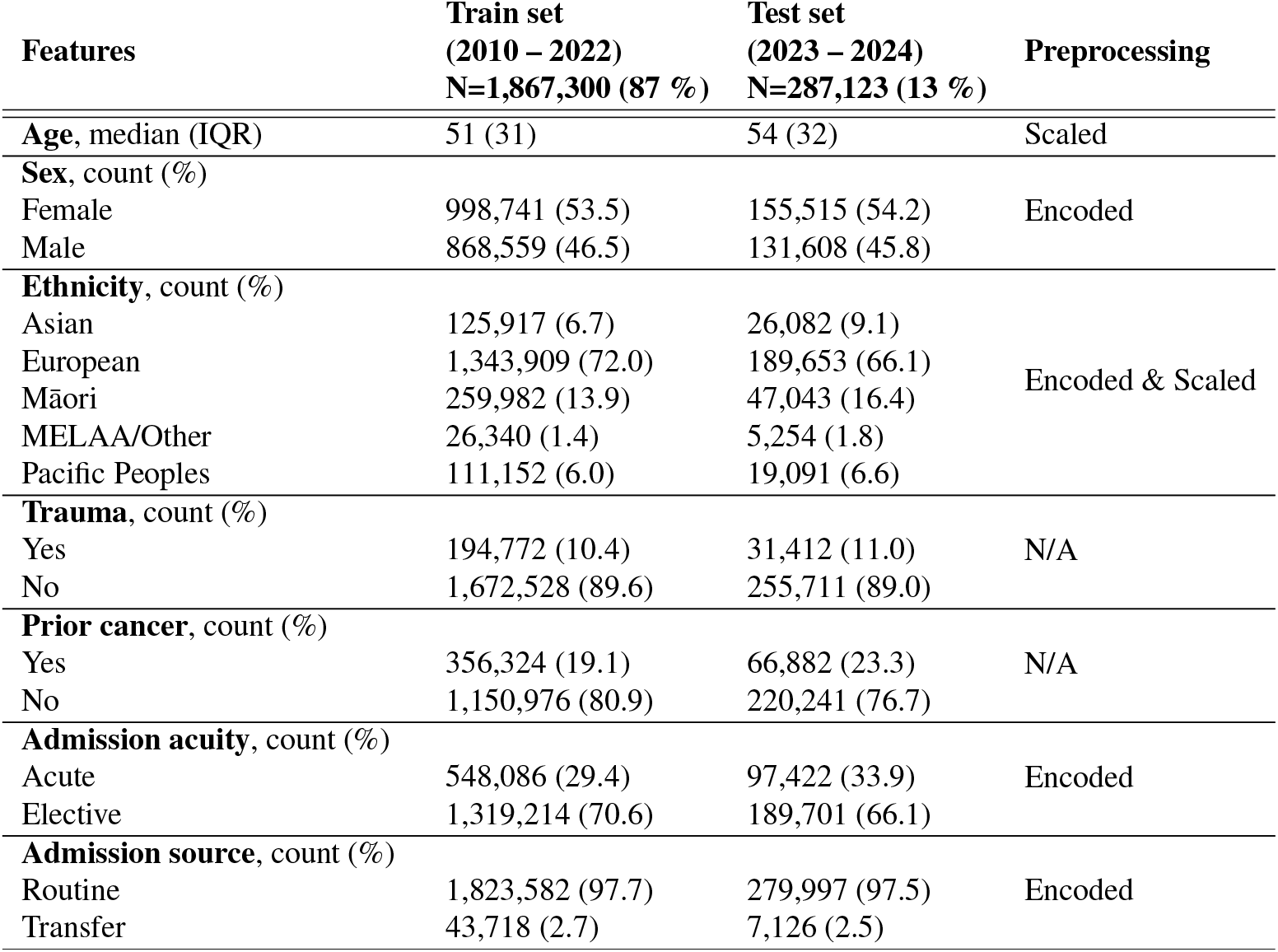
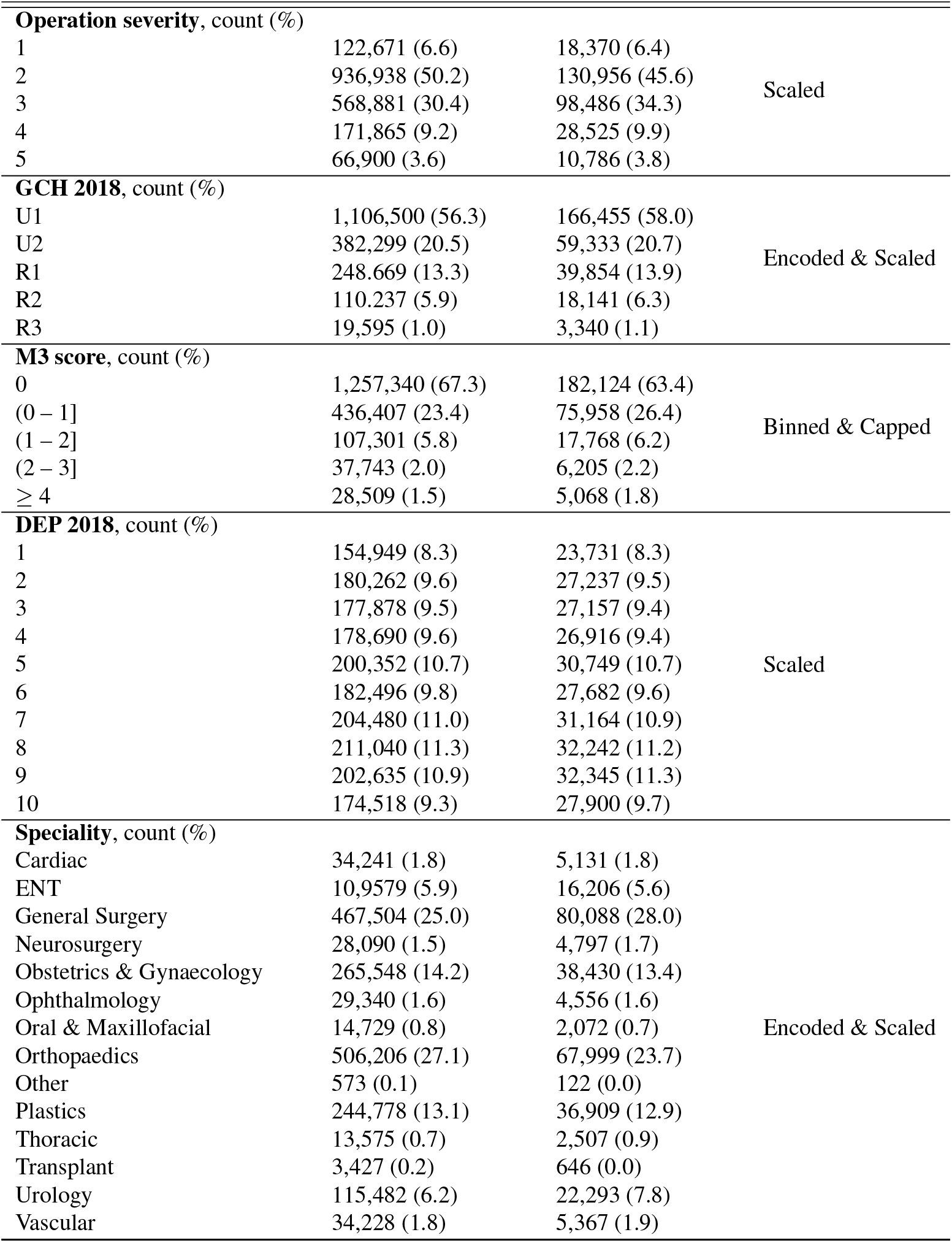
Participant characteristics and preprocessing methods.

## DISCUSSION

This paper presents the impacts of class imbalance mitigation strategies in AI prediction models for rare outcomes. We demonstrate substantial model miscalibration when class imbalance mitigation methods were applied, resulting in significant potential impacts to clinical decision-making.

In other AI domains, addressing class imbalance has been shown to significantly enhance predictive performance, possibly encouraging wider implementation of these methods. Notably, however, the improved performance metrics are usually based on threshold-dependent evaluation metrics, such as recall and F_1_ score. These metrics are typically prioritised to ensure models remains sensitive to high-risk patients ^23^, but, this apparent improvement in model performance is likely artefactual because class imbalance mitigation optimizes the learning problem toward binary separation under an implicitly balanced class prior. Metrics such as accuracy, recall, and F_1_ score require converting predicted probabilities into class labels, and their values can change markedly with the choice of threshold. When resampling (ROS/RUS/SMOTE) or aggressive class-weighting is applied, the effective prevalence seen during training is shifted upward toward 50%, which encourages the model to allocate more probability mass to the minority class. As a result, a conventional decision threshold of 0.5 becomes less stringent than it would be under the natural (low-prevalence) setting, so more cases are labelled “positive”. This typically increases sensitivity (recall) and can inflate F_1_, while reducing precision and often lowering overall accuracy—consistent with our findings.

At deployment, models are exposed to the natural distribution of risk in the real-world. Similarly, especially in healthcare settings, users prioritise well-calibrated probabilities under the natural outcome prevalence, where event rates may be very low. Artificially altering the class distribution can decouple model outputs from the true baseline risk, producing systematic shifts in the predicted probability distribution and, consequent output miscalibration. In clinical settings, where decision-making relies on precise risk stratification rather than simple binary labels, such miscalibration is a critical concern ^24;25;26^. Here we show that this can have significant impacts on downstream clinical decisions with proportion of surgeries classified as “high-risk” increasing by up to 62.8%, depending on the class imbalance mitigation method employed (Table 3). Such drastic variations highlight how artifactual shifts in probability can lead to over-diagnosis or the misallocation of clinical resources, underscoring the necessity of rigorous calibration in medical risk models. These results were supported by the decision analysis curves which showed that the natural model demonstrated a highest net benefit over a range of clinically relevant thresholds compared to all of the models using class imbalance mitigation strategies.

Van Calster *et al*. ^1^ showed that at clinically relevant thresholds, all classification measures (accuracy, recall, precision and F_1_ score) are improper, and recommend against their use. They define an improper measure as one that can lead to selecting an incorrect model because it outperforms the correct one. This is evident with the models developed here: the classification metrics were better for models that addressed class imbalance, however, their calibration was worse than the natural model trained on the natural data distribution (Figure 1). This result held regardless of the imbalance ratio value and the predicted outcome (Figure 3). The three other metrics (AUROC, AUPRC, and log loss), however, are threshold-independent and therefore fairer evaluations for comparing model discrimination and calibration. Therefore, model selection and reporting should emphasize on calibration curves, DCA, and threshold-independent discrimination metrics (*e*.*g*., AUROC, AUPRC, log loss). The latter measures evaluate performance across the full range of thresholds and penalize distorted probability estimates, making them more aligned with the objectives of clinical decision support than threshold-dependent classification metrics which are sensitive to prevalence and the chosen operating point. This is consistent with the latest reporting recommendations for AI predictive models ^1;18^.

A recent review of clinical prediction models developed using machine learning techniques found that 94.6% of models did not report calibration metrics and 17.8% used a class imbalance mitigation method ^27^. Previous simulation studies in synthetic datasets have highlighted the harms of class imbalance correction strategies, particularly resulting in miscalibrated model outputs ^24^. Other studies evaluated the effects of class imbalance in smaller datasets ^28;26^ and found similar results with comparable discrimination and potentially harmful effects on calibration. We extend these results by evaluating the impact of various degrees of class imbalance correction across a range of imbalance ratios on discrimination and calibration for multiple outcomes in a much larger dataset.

These findings reflect the importance of the pre-test probability in clinical prediction models. As per Bayesian principles, all clinical prediction model outputs should be understood as a posterior probability that integrates prior information with patient-specific data ^29^. When class imbalance mitigation strategies artificially alter the pre-test probability distribution seen during model training, they effectively recast the population context to which the risk estimates of the model apply, explaining the arising miscalibration ^28^. This disconnect between training distribution and real-world prevalence compromises the interpretability and clinical validity of risk estimates, particularly when decisions depend on specific risk thresholds, which may vary based on clinical contexts. Post-hoc calibration methods, such as Platt scaling or isotonic regression, can be used to recalibrate the probabilities of a model and improve the calibration of the model ^30^. Recalibrating the models that used class imbalance mitigation methods led to a much better calibration (see Supplemental Figure 7). However, the probability distribution was similar to the natural distribution, which puts into question the need for addressing class imbalance.

This study is not without limitations. The HGB classifier is highly effective with tabular data, however, other models that are used clinically such logistic regressions ^26^, or deep neural networks ^31;32^ may yield different results. Consequently, our findings regarding the natural distribution of events may vary when applied to models with different inductive biases. The large-scale dataset that was used provided the statistical power necessary to observe subtle calibration drifts. However, the trade-offs observed here may differ in smaller cohorts where insufficient minority class representation poses a greater risk of model under-fitting. Finally, this work focuses on clinical scenarios where output probabilities are important. Class imbalance mitigation may remain a valuable tool in settings where only the rank-order or binary identification is required.

## CONCLUSION

This study demonstrated in a large-scale clinical dataset, that class imbalance mitigation strategies for predicting rare outcomes resulted in demonstrable model miscalibration and possible clinical harm. In our analysis, the model trained on the natural, imbalanced distribution consistently outperformed mitigated models in decision-making utility. These findings suggest that in clinical environments, the accuracy of a predicted probabilities (*i*.*e*., calibration) is more critical than the maximization of binary classification metrics.

## DATA AVAILABILITY

Data can be obtained from the New Zealand Ministry of Health National Collections team. Requests can be made by contacting. A Python package was created to create training and evaluation pipelines and is freely available on Github at <https://github.com/Surgical-Recovery-and-Safety-Lab/medpipe>. The code used to train the models and create the figures is freely available on GitHub at https://github.com/Surgical-Recovery-and-Safety-Lab/AI-risk-score.

## AUTHOR CONTRIBUTIONS

**Mathias Roesler**: Conceptualisation, Methodology, Software, Formal Analysis, Writing - Original Draft. **Cameron Wells**: Data Curation, Writing - Review & Editing. **Gabriel Schamberg**: Writing - Review & Editing. **Junyi Gao**: Writing - Review & Editing. **Luke Boyle**: Data Curation, Writing - Review & Editing. **Greg O’Grady**: Writing - Review & Editing. **Chris Varghese**: Conceptualisation, Methodology, Formal Analysis, Writing - Original Draft.

## ACKNOWLEDGMENTS

This work was funded by the Health Research Council of New Zealand 2025 Artificial Intelligence (AI) in Healthcare Project Grant.

## SUPPLEMENTAL MATERIALS

Figure 7 shows the calibration curves and predicted probability distributions of the models that used class imbalance mitigation methods before and after post-hoc calibration. The recalibration was performed using an isotonic regression model and the test data from the year 2023 and the plots were generated from the test data from the year 2024, thereby ensuring proper external validation. The probability distributions were plotted with a log scale on the y-axis.

**Figure 7:**
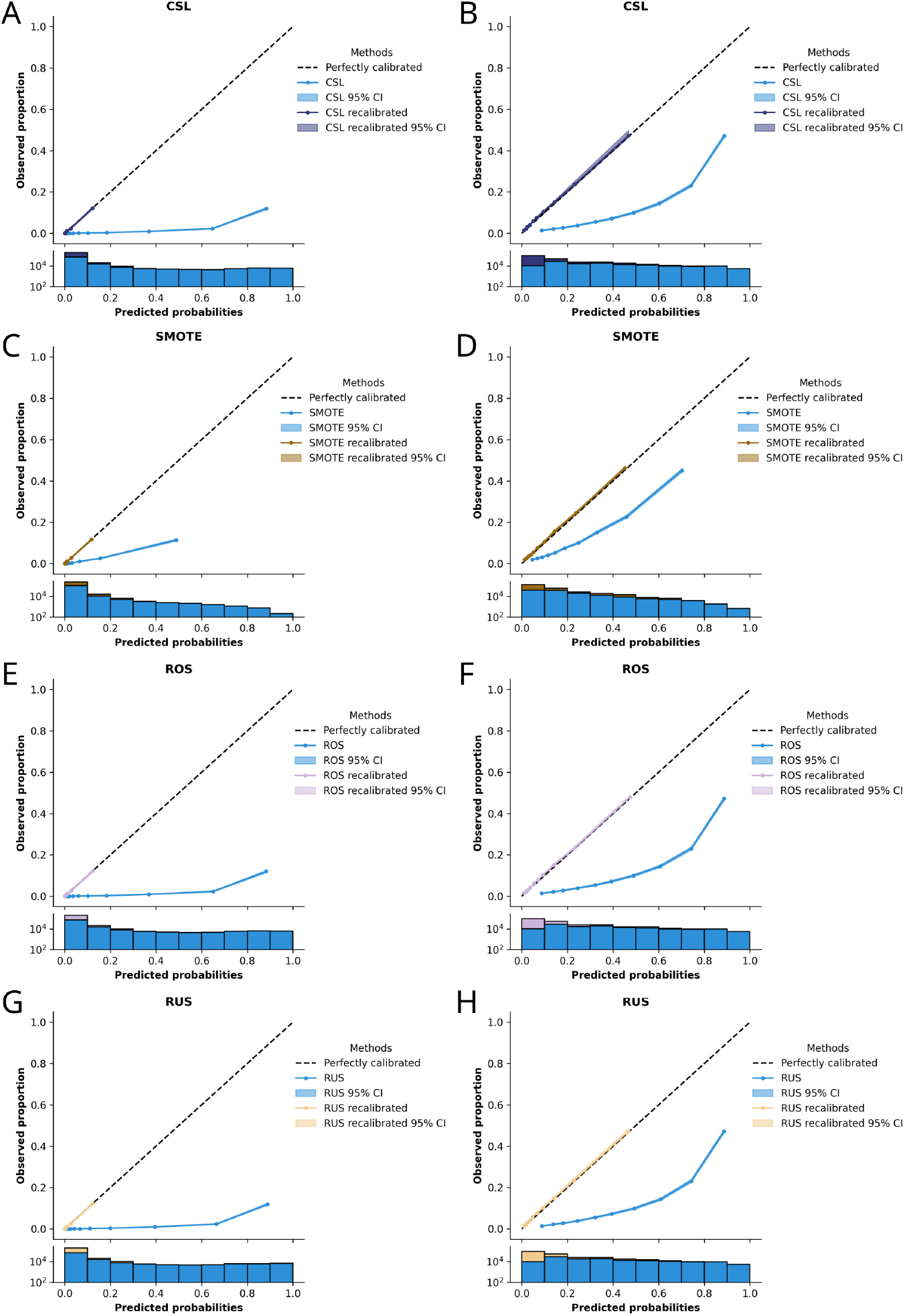
Post-hoc calibration of the models using class imbalance mitigation methods and an imbalance ratio (IR) of 1. The graphs on the left (panels A, C, and E) show the calibration curves for the 90-day mortality label while those on the right (panels B, D, and F) correspond to the complications label. The calibration curves followed the perfect calibration line much closer after recalibration. The probability distributions resembled the natural distribution, which a much higher incidence rate between 0 and 0.1.

