## Supplemental Materials for "Class imbalance correction in artificial intelligence models leads to miscalibrated clinical predictions: a real-world evaluation"

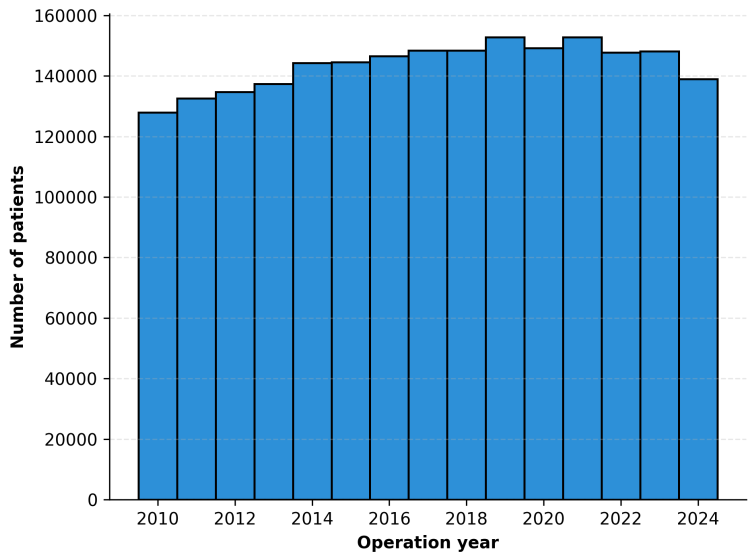


**Figure 6:** Number of patients per operation year. The distribution showed that the number of patients increased between 2010 and 2014 but remained stable afterwards. This distribution confirms the use of a time-based cross-validation training scheme.

**Table 4:** Participant characteristics and preprocessing methods.

| **Features** | **Train set (2010 – 2022) N=1,867,300 (87 %)** | **Test set (2023 – 2024) N=287,123 (13 %)** | **Preprocessing** |
|---|---|---|---|
| **Age**, median (IQR) | 51 (31) | 54 (32) | Scaled |
| **Sex**, count (%) | | | |
| Female | 998,741 (53.5) | 155,515 (54.2) | Encoded |
| Male | 868,559 (46.5) | 131,608 (45.8) | |
| **Ethnicity**, count (%) | | | |
| Asian | 125,917 (6.7) | 26,082 (9.1) | |
| European | 1,343,909 (72.0) | 189,653 (66.1) | Encoded & Scaled |
| Māori | 259,982 (13.9) | 47,043 (16.4) | |
| MELAA/Other | 26,340 (1.4) | 5,254 (1.8) | |
| Pacific Peoples | 111,152 (6.0) | 19,091 (6.6) | |
| **Trauma**, count (%) | | | |
| Yes | 194,772 (10.4) | 31,412 (11.0) | N/A |
| No | 1,672,528 (89.6) | 255,711 (89.0) | |
| **Prior cancer**, count (%) | | | |
| Yes | 356,324 (19.1) | 66,882 (23.3) | N/A |
| No | 1,150,976 (80.9) | 220,241 (76.7) | |
| **Admission acuity**, count (%) | | | |
| Acute | 548,086 (29.4) | 97,422 (33.9) | Encoded |
| Elective | 1,319,214 (70.6) | 189,701 (66.1) | |
| **Admission source**, count (%) | | | |
| Routine | 1,823,582 (97.7) | 279,997 (97.5) | Encoded |
| Transfer | 43,718 (2.7) | 7,126 (2.5) | |

*Continued on next page*

| Features | Train set (2010–2022) N=1,867,300 (87 %) | Test set (2023 – 2024) N=287,123 (13 %) | Preprocessing |
|---|---|---|---|
| **Operation severity**, count (%) | | | |
| 1 | 122,671 (6.6) | 18,370 (6.4) | |
| 2 | 936,938 (50.2) | 130,956 (45.6) | Scaled |
| 3 | 568,881 (30.4) | 98,486 (34.3) | |
| 4 | 171,865 (9.2) | 28,525 (9.9) | |
| 5 | 66,900 (3.6) | 10,786 (3.8) | |
| **GCH 2018**, count (%) | | | |
| U1 | 1,106,500 (56.3) | 166,455 (58.0) | |
| U2 | 382,299 (20.5) | 59,333 (20.7) | Encoded & Scaled |
| R1 | 248.669 (13.3) | 39,854 (13.9) | |
| R2 | 110.237 (5.9) | 18,141 (6.3) | |
| R3 | 19,595 (1.0) | 3,340 (1.1) | |
| **M3 score**, count (%) | | | |
| 0 | 1,257,340 (67.3) | 182,124 (63.4) | |
| (0 – 1] | 436,407 (23.4) | 75,958 (26.4) | Binned & Capped |
| (1 – 2] | 107,301 (5.8) | 17,768 (6.2) | |
| (2 – 3] | 37,743 (2.0) | 6,205 (2.2) | |
| $\geq 4$ | 28,509 (1.5) | 5,068 (1.8) | |
| **DEP 2018**, count (%) | | | |
| 1 | 154,949 (8.3) | 23,731 (8.3) | |
| 2 | 180,262 (9.6) | 27,237 (9.5) | |
| 3 | 177,878 (9.5) | 27,157 (9.4) | |
| 4 | 178,690 (9.6) | 26,916 (9.4) | |
| 5 | 200,352 (10.7) | 30,749 (10.7) | Scaled |
| 6 | 182,496 (9.8) | 27,682 (9.6) | |
| 7 | 204,480 (11.0) | 31,164 (10.9) | |
| 8 | 211,040 (11.3) | 32,242 (11.2) | |
| 9 | 202,635 (10.9) | 32,345 (11.3) | |
| 10 | 174,518 (9.3) | 27,900 (9.7) | |
| **Speciality**, count (%) | | | |
| Cardiac | 34,241 (1.8) | 5,131 (1.8) | |
| ENT | 10,9579 (5.9) | 16,206 (5.6) | |
| General Surgery | 467,504 (25.0) | 80,088 (28.0) | |
| Neurosurgery | 28,090 (1.5) | 4,797 (1.7) | |
| Obstetrics & Gynaecology | 265,548 (14.2) | 38,430 (13.4) | |
| Ophthalmology | 29,340 (1.6) | 4,556 (1.6) | |
| Oral & Maxillofacial | 14,729 (0.8) | 2,072 (0.7) | Encoded & Scaled |
| Orthopaedics | 506,206 (27.1) | 67,999 (23.7) | |
| Other | 573 (0.1) | 122 (0.0) | |
| Plastics | 244,778 (13.1) | 36,909 (12.9) | |
| Thoracic | 13,575 (0.7) | 2,507 (0.9) | |
| Transplant | 3,427 (0.2) | 646 (0.0) | |
| Urology | 115,482 (6.2) | 22,293 (7.8) | |
| Vascular | 34,228 (1.8) | 5,367 (1.9) | |

Figure 7 shows the calibration curves and predicted probability distributions of the models that used class imbalance mitigation methods before and after post-hoc calibration. The recalibration was performed using an isotonic regression model and the test data from the year 2023 and the plots were generated from the test data from the year 2024, thereby ensuring proper external validation. The probability distributions were plotted with a log scale on the y-axis.

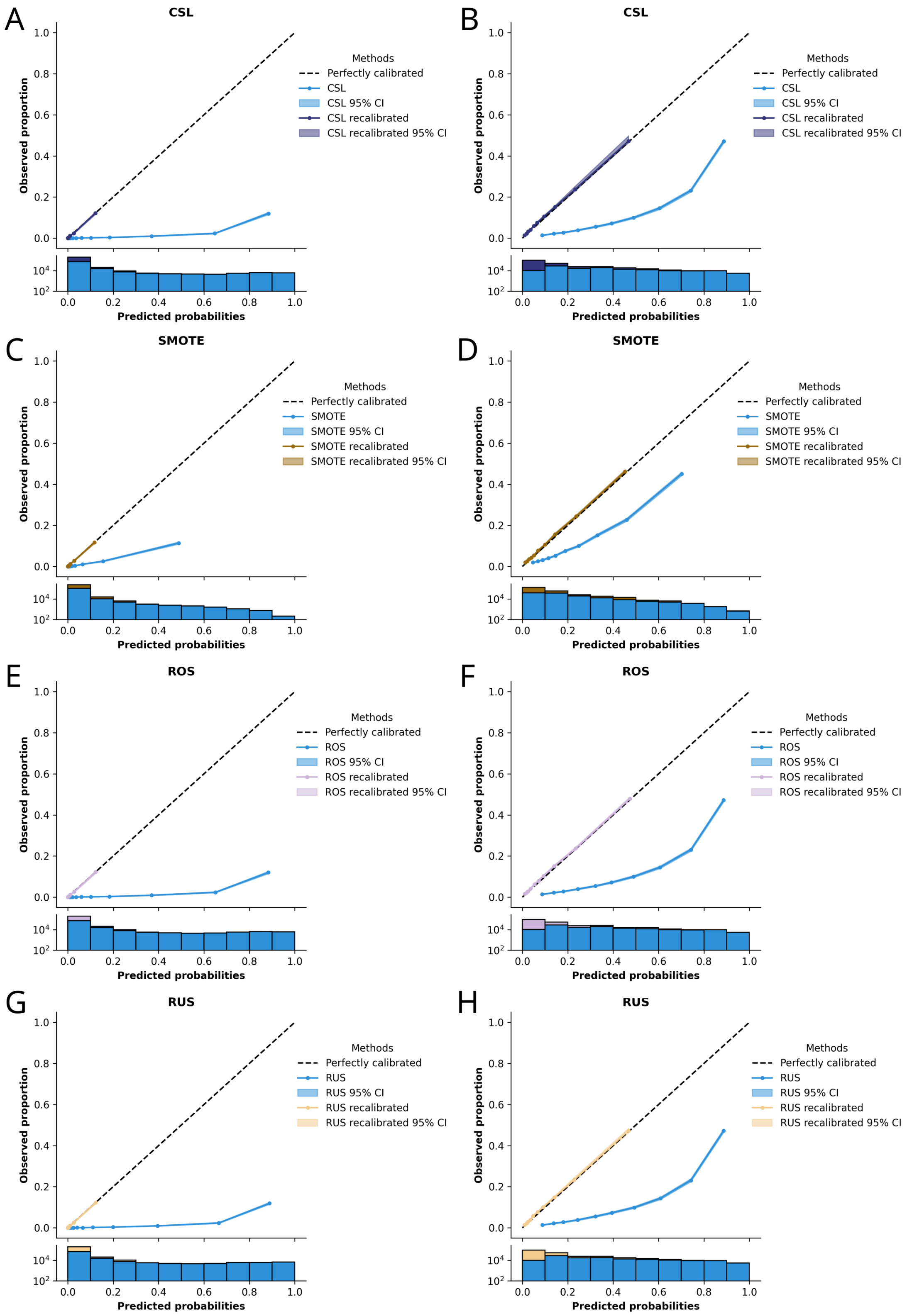


**Figure 7:** Post-hoc calibration of the models using class imbalance mitigation methods and an imbalance ratio (IR) of 1. The graphs on the left (panels A, C, and E) show the calibration curves for the 90-day mortality label while those on the right (panels B, D, and F) correspond to the complications label. The calibration curves followed the perfect calibration line much closer after recalibration. The probability distributions resembled the natural distribution, which a much higher incidence rate between 0 and 0.1.
